# Risk of COVID-19 among frontline healthcare workers and the general community: a prospective cohort study

**DOI:** 10.1101/2020.04.29.20084111

**Authors:** Long H. Nguyen, David A. Drew, Amit D. Joshi, Chuan-Guo Guo, Wenjie Ma, Raaj S. Mehta, Daniel R. Sikavi, Chun-Han Lo, Sohee Kwon, Mingyang Song, Lorelei A. Mucci, Meir J. Stampfer, Walter C. Willett, A. Heather Eliassen, Jaime E. Hart, Jorge E. Chavarro, Janet W. Rich-Edwards, Richard Davies, Joan Capdevila, Karla A. Lee, Mary Ni Lochlainn, Thomas Varsavsky, Mark S. Graham, Carole H. Sudre, M. Jorge Cardoso, Jonathan Wolf, Sebastien Ourselin, Claire J. Steves, Tim D. Spector, Andrew T. Chan, On behalf of the COPE Consortium

## Abstract

**Background:** Data for frontline healthcare workers (HCWs) and risk of SARS-CoV-2 infection are limited and whether personal protective equipment (PPE) mitigates this risk is unknown. We evaluated risk for COVID-19 among frontline HCWs compared to the general community and the influence of PPE.

**Methods:** We performed a prospective cohort study of the general community, including frontline HCWs, who reported information through the COVID Symptom Study smartphone application beginning on March 24 (United Kingdom, U.K.) and March 29 (United States, U.S.) through April 23, 2020. We used Cox proportional hazards modeling to estimate multivariate-adjusted hazard ratios (aHRs) of a positive COVID-19 test.

**Findings:** Among 2,035,395 community individuals and 99,795 frontline HCWs, we documented 5,545 incident reports of a positive COVID-19 test over 34,435,272 person-days. Compared with the general community, frontline HCWs had an aHR of 11·6 (95% CI: 10·9 to 12·3) for reporting a positive test. The corresponding aHR was 3·40 (95% CI: 3·37 to 3·43) using an inverse probability weighted Cox model adjusting for the likelihood of receiving a test. A symptom-based classifier of predicted COVID-19 yielded similar risk estimates. Compared with HCWs reporting adequate PPE, the aHRs for reporting a positive test were 1·46 (95% CI: 1·21 to 1·76) for those reporting PPE reuse and 1·31 (95% CI: 1·10 to 1·56) for reporting inadequate PPE. Compared with HCWs reporting adequate PPE who did not care for COVID-19 patients, HCWs caring for patients with documented COVID-19 had aHRs for a positive test of 4·83 (95% CI: 3·99 to 5·85) if they had adequate PPE, 5·06 (95% CI: 3·90 to 6·57) for reused PPE, and 5·91 (95% CI: 4·53 to 7·71) for inadequate PPE.

**Interpretation:** Frontline HCWs had a significantly increased risk of COVID-19 infection, highest among HCWs who reused PPE or had inadequate access to PPE. However, adequate supplies of PPE did not completely mitigate high-risk exposures.

**Funding:** Zoe Global Ltd., Wellcome Trust, EPSRC, NIHR, UK Research and Innovation, Alzheimer’s Society, NIH, NIOSH, Massachusetts Consortium on Pathogen Readiness

**RESEARCH IN CONTEXT:** *Evidence before this study:* The prolonged course of the coronavirus disease 2019 (COVID-19) pandemic, coupled with sustained challenges supplying adequate personal protective equipment (PPE) for frontline healthcare workers (HCW), have strained global healthcare systems in an unprecedented fashion. Despite growing awareness of this problem, there are few data to inform policy makers on the risk of COVID-19 among HCWs and the impact of PPE on their disease burden. Prior reports of HCW infections are based on cross sectional data with limited individual-level information on risk factors for infection. A PubMed search for articles published between January 1, 2020 and May 5, 2020 using the terms “covid-19”, “healthcare workers”, and “personal protective equipment,” yielded no population-scale investigations exploring this topic.

*Added value of this study:* In a prospective study of 2,135,190 individuals, frontline HCWs may have up to a 12-fold increased risk of reporting a positive COVID-19 test. Compared with those who reported adequate availability of PPE, frontline HCWs with inadequate PPE had a 31% increase in risk. However, adequate availability of PPE did not completely reduce risk among HCWs caring for COVID-19 patients.

*Implications of all the available evidence:* Beyond ensuring adequate availability of PPE, additional efforts to protect HCWs from COVID-19 are needed, particularly as lockdown is lifted in many regions of the world.

## INTRODUCTION

Since its emergence, SARS-CoV-2 has become a global health threat.^1^ As of May 2020, over 3·8 million cases of COVID-19 have been documented worldwide with nearly 270,000 deaths.^2^ With ongoing community transmission from asymptomatic individuals, the burden of this disease is expected to rise over the coming weeks. Consequently, there will be an ongoing need for frontline healthcare workers (HCW) in patient-facing roles.^3^ Because this work requires close personal exposure to patients with the virus, frontline HCWs are at high risk of infection, which may contribute to further spread.^4^ Initial regional estimates suggest frontline HCWs may account for 10-20% of all diagnoses,^5-7^ which may be an underestimate when compared to other developed and similarly affected nations such as Italy.^8^

Based on experience with other viruses spread by respiratory droplets, the consistent use of recommended personal protective equipment (PPE) is critical to reducing nosocomial transmission.^9^ Recent guidelines from the United States (U.S.) Center for Disease Control and Prevention (CDC) recommend respirator use when caring for patients with suspected or confirmed COVID-19 and the universal use of masks at work.^10^ Joint guidelines from governing health bodies in Europe, including the National Health Service in the United Kingdom (U.K.), issued similarly graduated PPE recommendations dictated by the intensity of clinical exposure and likelihood of contact with bodily secretions.^11^ However, global shortages of masks, face shields, and gowns—caused by surging demand and supply chain disruptions—have been documented, leading to efforts to conserve PPE through extended use or reuse and the recent development of disinfection protocols for which there remains a lack of peer-reviewed, scientific consensus on best practices.^12-14^

Although addressing the needs of frontline HCWs to respond to the COVID-19 pandemic is a high priority,^3,7^ there is a lack of data to inform such efforts. Thus, we conducted a prospective, population-based study using a novel mobile-based application to examine the risk of testing positive for COVID-19 and/or developing symptoms associated with infection among individuals in the U.K. and the U.S. between March 24 and April 23, 2020.

## METHODS

### Development and deployment of a smartphone application

This prospective observational cohort study was conducted using the COVID Symptom Study (previously known as the COVID Symptom Tracker) app, a freely available smartphone application developed by Zoe Global Ltd. in collaboration with the Massachusetts General Hospital and King’s College London that offers participants a guided interface to report a range of baseline demographic information and comorbidities, daily information on potential symptoms, and COVID-19 testing. Participants are encouraged to use the application daily, even when asymptomatic, to allow for the longitudinal, prospective collection of symptoms and COVID-19 testing results.

### Study design and participants

The application was launched in the U.K. on March 24, 2020 and available in the U.S. beginning on March 29, 2020. Participants were recruited through social media outreach, as well as invitations from the investigators of long-running cohort studies to study volunteers (**Suppl. Table 1**). At enrollment, participants provided informed consent to the use of aggregated information for research purposes and agreed to applicable privacy policies and terms of use. This observational study was approved by the Partners Human Research Committee (Protocol 2020P000909) and King’s College London Ethics Committee (REMAS ID 18210, LRS-19/20-18210). This protocol is registered with ClinicalTrials.gov (NCT04331509).

### Assessment of risk factors, symptoms, and testing

Information collected through the app has been provided in detail.^15^ Briefly, upon first use, participants were asked to provide demographic factors and answered separate questions about a series of suspected risk factors for COVID-19 (**Suppl. Table 2** and **Suppl. Table 3**). At enrollment and upon daily reminders, participants were asked if they felt physically normal, and if not, their symptoms (**Suppl. Table 4**). Participants were also asked if they had been tested for COVID-19 (yes/no), and if yes, the results (none, negative, pending, or positive).

At enrollment, individuals were asked if they worked in health care and if yes, whether they had direct patient contact. For our primary analysis, we defined frontline HCWs as participants who reported direct patient contact. Among these individuals, we queried whether they cared for suspected or documented COVID-19-infected patients and the frequency with which they used PPE (always, sometimes, never). We asked if they had enough PPE when needed, if they had to reuse PPE, or if they did not have enough because of shortages. We classified availability of PPE as adequate if they never required PPE or if they reported always having the PPE they needed. We classified PPE availability as inadequate if they reported they did not have enough PPE or if it was not available. We also asked HCWs to report the site of their patient care.

### Statistical Analysis

Follow-up time started when participants first reported on the app and accrued until the report of a positive COVID-19 test or the time of last data entry, whichever occurred first. We employed Cox proportional hazards modeling stratified by age, date, and country to estimate age- and multivariable-adjusted hazard ratios (aHRs) and their 95% confidence intervals (95% CIs). A test of correlation between Schoenfeld residuals and survival time demonstrated no violation of the proportional hazards assumption. Covariates were selected *a priori* based on putative risk factors and included sex, history of diabetes, heart disease, lung disease, kidney disease, current smoking status (each yes/no), and body mass index (17-19·9, 20-24·9, 25-29·9, and ≥30 kg/m^2^). Data imputation replaced no more than 5% of missing values for a given metadatum. Missing numeric values were replaced with the median value, while categorical variables were imputed using the mode.

Because the outcome for our primary analysis (report of a positive COVID-19 test) required receiving a test, we performed several secondary analyses to ensure the robustness of our findings. First, we leveraged a symptom-based classifier developed by our group that is predictive of positive COVID-19 testing.^16^ Briefly, using logistic regression and symptoms preceding confirmatory testing, we found that loss of smell/taste, fatigue, persistent cough, and loss of appetite predicts COVID-19 positivity with high specificity (**Suppl. Methods**). Second, to account for country-specific predictors of obtaining testing, we performed separate inverse probability weighting (IPW) in the U.S. and the U.K. as a function of demographic and clinical factors, such as age and symptom burden, followed by inverse probability weighted-Cox proportional hazards modeling stratified by 5-year age group and date with additional adjustment for the covariates used in prior models (**Suppl. Methods**). In analyses limited to frontline HCWs, we examined PPE availability and contact with suspected or documented COVID-19 patients, as well as the primary site of clinical practice. Two-sided *p*-values <0·05 were considered statistically significant. All analyses were performed using R 3·6·1 (Vienna, Austria).

## RESULTS

### Study population

Between March 24 and April 23, 2020, we enrolled 2,810,103 users (2,627,695 in the U.K. and 182,408 in the U.S.), defined as participants who provided baseline information about either feeling normal or having symptoms (**Suppl. Fig. 1**). Among users, 134,885 (4·8%) reported being a frontline HCW. We found a reported prevalence of 2,747 COVID-19 cases per 100,000 frontline HCWs compared to 242 per 100,000 in the general community (**Fig. 1A**). Higher infection rates were reported in New York, New Jersey, and Louisiana in the U.S. and in the areas around London and the Midlands in the U.K. (**Fig. 1B**).

**Figure 1.**
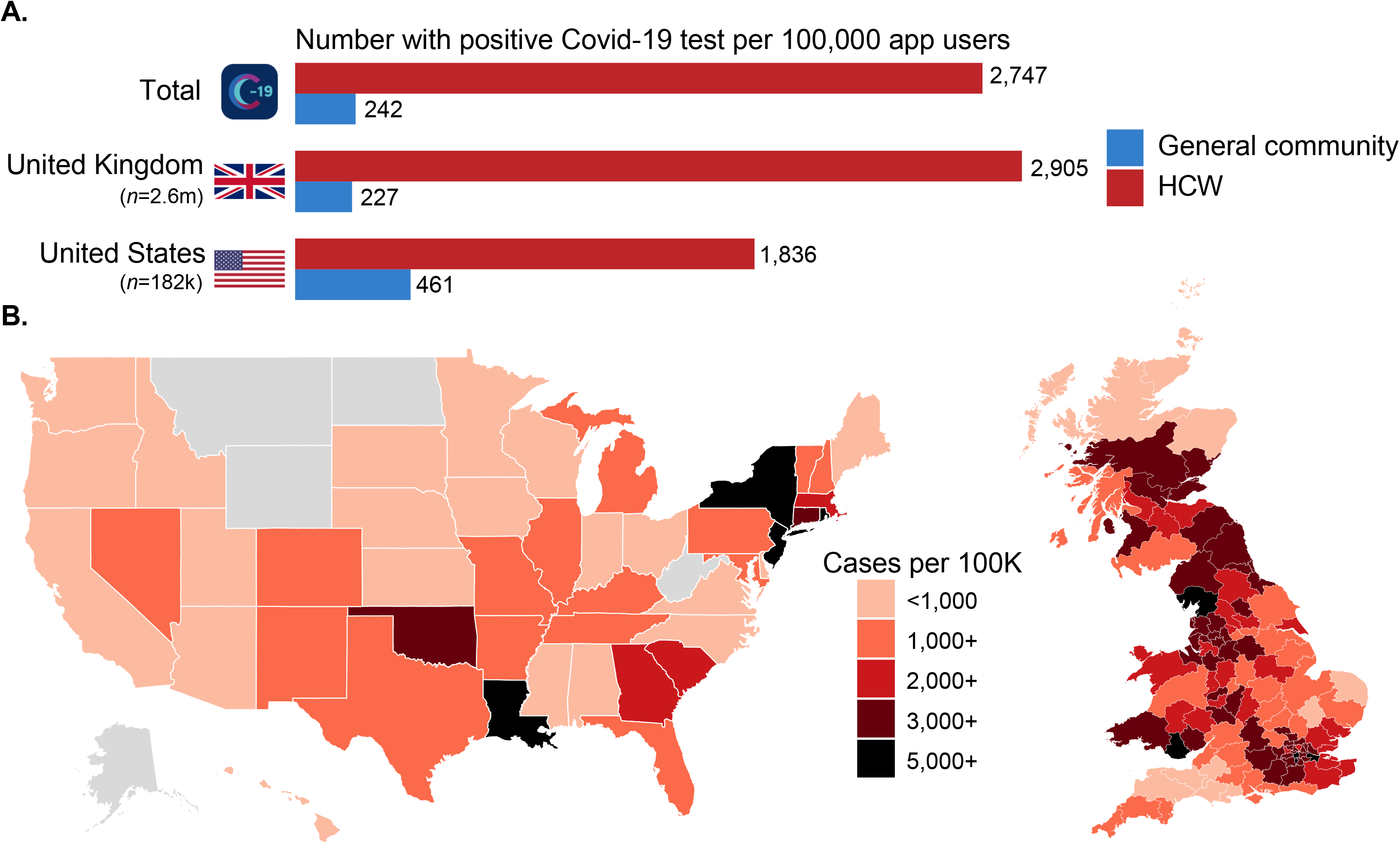
The risk of testing positive for COVID-19 among frontline healthcare workers (HCW) **A**. Between March 24, 2020 and April 23, 2020, considerable disparities in prevalence of a positive COVID-19 test among frontline HCW risk compared to the general community were observed in both the United Kingdom and the United States. **B**. Prevalence of a positive COVID-19 test reported by frontline HCWs in the United States and the United Kingdom. Regions in gray did not offer sufficient data.

After excluding 670,298 participants who had follow-up time of less than 24 hours and 4,615 who reported a baseline positive COVID-19 test, we included 2,135,190 participants in our prospective inception cohort, among whom 99,795 (4·7%) persons identified as frontline HCWs (**Suppl. Fig. 1**). In this cohort, we recorded 24·4 million entries or 11·5 logs per participant with a median follow-up of 18·9 days (interquartile range [IQR] 5·1 to 26·1). The median age was 44 years (IQR 32 to 57). Compared to the general community, frontline HCWs were more frequently female, had a slightly higher prevalence of BMI ≥30, were slightly more likely to smoke (particularly in the U.K.), and use several common medications (**Table 1** and **Suppl. Table 3**). At baseline, 20·2% of frontline HCWs reported at least one symptom associated with COVID-19 infection compared to 14·4% of the general population with fatigue, loss of smell/taste, and hoarse voice being particularly more frequent (**Suppl. Table 4**).

**Table 1.** Baseline characteristics of frontline healthcare workers compared to the general public.

|  | Participants<br>(*n*=2,135,190) |  |
| --- | --- | --- |
|  | Frontline HCWs<br>(*n*=99,795) | General community<br>(*n*=2,035,395) |
| **Country (%)** |  |  |
| U.S. | 14.6 | 6.1 |
| U.K. | 85.4 | 93.9 |
| **Age (years)** | 42 [33, 53] | 44 [33, 56] |
| <25 | 4.5 | 5.0 |
| 25-34 | 24.7 | 20.4 |
| 35-44 | 25.4 | 22.8 |
| 45-54 | 33.9 | 20.7 |
| 55-64 | 17.7 | 17.2 |
| ≥65 | 3.9 | 13.9 |
| Missing | 1.1 | 5.7 |
| **Male sex (%)** | 17.0 | 37.0 |
| **Race (%)** |  |  |
| Black | 1.9 | 1.4 |
| White | 92.2 | 94.6 |
| Asian | 4.2 | 2.1 |
| Other | 1.7 | 1.8 |
| Missing/Prefer not to say | 5.0 | 2.0 |
| **BMI (kg/m<sup>2</sup>)** | 25.8 [22.8, 30.2] | 25.3 [22.5, 29.1] |
| 17-19.9 | 5.8 | 8.3 |
| 20-24.9 | 38.1 | 39.2 |
| 25-29.9 | 30.1 | 31.5 |
| ≥30 | 25.9 | 21.1 |
| Missing | 0.5 | 0.5 |
| **Comorbidities (%)** |  |  |
| Diabetes | 2.5 | 3.1 |

|  | **Participants**<br>**(n=2,135,190)** |  |
| --- | --- | --- |
|  | **Frontline HCWs**<br>**(n=99,795)** | **General community**<br>**(n=2,035,395)** |
| Heart Disease | 1.6 | 2.4 |
| Lung Disease | 13.1 | 12.2 |
| Kidney Disease | 0.6 | 0.7 |
| Cancer |  |  |
| Yes | 0.5 | 1.3 |
| Missing | 0.3 | 0.3 |
| **Pregnant (% of females)** | 0.9 | 1.0 |
| **Medication usage (%)** |  |  |
| NSAIDs | 8.2 | 6.1 |
| Immunosuppressants | 2.5 | 3.2 |
| Chemotherapy/Immunotherapy | 0.1 | 0.3 |
| ACE inhibitor | 5.0 | 4.9 |
| Yes | 4.5 | 4.6 |
| Missing | 10.1 | 4.3 |
| **Current smoking (%)** | 9.7 | 7.6 |
| Yes | 10.2 | 8.5 |
| Missing | 0.2 | 0.1 |

Abbreviations: ACE (angiotensin converting enzyme), BMI (body mass index), m (meter), kg (kilogram),
Non-steroidal anti-inflammatory drugs (NSAIDs)
Median [IQR] is presented for continuous variables. Frequencies and proportions are calculated based on the total number of participants with available data.
History of cancer, ACE inhibitor use, and smoking status have been queried since launch in the U.S. and since 3/29/2020 in the U.K. Race was queried as of 4/17/2020.
Definitions for race: Black (U.K. Black/Black British, U.K. Mixed Race-White and Black/Black British, U.S. Black or African-American), White (U.K. White, U.S. White), Asian (U.K. Asian/Asian British, U.K. Chinese/Chinese British, U.S. Asian, U.S. Native Hawaiian or Other Pacific Islander), and Other (U.K. Mixed Race Other, U.K. Middle Eastern/Middle Eastern British, U.S. American Indian or Alaska Native, Other).

### Risk of positive COVID-19 testing and symptoms in HCWs

We documented 5,545 incident reports of positive COVID-19 testing over 34,435,272 person-days. In the U.K, 1.1% of HCWs reported being tested for COVID-19 compared to 0.2% of the general community (ratio of testing HCWs/community testing: 5.5), while 4.1% of U.S. HCWs were tested vs. 1.1% of the general community (ratio: 3.7). Compared with the general community, frontline HCWs had a 12-fold increase in risk of a positive test after multivariable adjustment (aHR 11·6, 95% CI: 10·9 to 12·3; **Table 2** and **Suppl. Fig. 2**). Notably, the association appeared stronger in the U.K. (aHR 12·5, 95% CI: 11·8 to 13·3) compared to the U.S. (aHR 2·87, 95% CI: 2·14 to 3·85, *p*_difference_<0·0001; **Suppl. Table 5**). An analysis according to sex demonstrated similar risk estimates among male (aHR 14·0, 95% CI: 12·4 to 15·8) and female (aHR 11·3, 95% CI: 10·5-12·1) frontline HCWs.

**Table 2.** Risk of reporting a positive test for COVID-19 or predicted COVID-19 infection among HCWs compared with the general community.

|  | No. with<br>Event/Person-days | Incidence<br>(30-day) | Hazard Ratio (95% CI) |  |  |
| --- | --- | --- | --- | --- | --- |
|  |  |  | Age-adjusted | Multivariate-adjusted | IP Weighted |
| **<u>Positive COVID-19 testing</u>** |  |  |  |  |  |
| General community | 3,623/32,980,571 | 0.33% | 1.0 (ref.) | 1.0 (ref.) | 1.0 (ref.) |
| Frontline healthcare worker | 1,922/1,454,701 | 3.96% | 11.7 (11.0 to 12.4) | 11.6 (10.9 to 12.3) | 3.40 (3.37 to 3.43) |
| **<u>Predicted COVID-19 infection<sup>a</sup></u>** |  |  |  |  |  |
| General community | 56,059/29,864,522 | 5.63% | 1.0 (ref.) | 1.0 (ref.) | NA |
| Frontline healthcare worker | 5,022/1,242,857 | 12.1% | 2.05 (1.99 to 2.11) | 2.04 (1.98 to 2.10) | NA |

Abbreviations: CI (confidence interval), IP (inverse probability)
All models were stratified by 5-year age group, calendar date at study entry, and country.
Multivariate risk factor models were adjusted for sex, history of diabetes, heart disease, lung disease, kidney disease, and current smoking (each yes/no), and body mass index (17-19.9, 20-24.9, 25-29.9, and $\geq 30$ kg/m<sup>2</sup>).
<sup>a</sup>Using a symptom-based model described in Menni et al (*Nature Medicine* 2020)

We considered the possibility that the observed difference in risk in the U.K. versus the U.S. may be related to differences in the risk profile of individuals eligible for testing. A multivariable-adjusted Cox proportional hazards model with inverse probability weighting for predictors of testing also demonstrated higher risk of infection among frontline HCWs (aHR 3·40, 95%: 3·37 to 3·43; Table 2) with higher observed risk in the U.K. (aHR 3·43, 95% CI: 3·18 to 3·69). compared with the U.S. (aHR 1·97, 95% CI: 1·36 to 2·85, *p*_difference_ < 0·0001; **Suppl. Table 5**). In a secondary analysis, we used a validated model based on a combination of symptoms to predict likely COVID-19 infection.^16^ Compared with the general community, HCWs initially free of symptoms had an aHR of 2·05 (95% CI: 1·99 to 2·10; **Table 2**) for predicted COVID-19 that was also higher in the U.K (aHR 2·09, 95% CI: 2·02 to 2·15) than in the U.S. (aHR 1·31, 95% CI: 1·14 to 1·51, *p*_difference_<0·0001; **Suppl. Table 5**).

### PPE usage in frontline HCWs

Among frontline HCWs, we assessed PPE in relation to COVID-19 patient exposures and subsequent risk for testing positive. Compared with HCWs endorsing adequate PPE, frontline HCWs reporting the reuse of PPE had a 46% increased risk of reporting a positive COVID-19 test (aHR 1·46, 95% CI: 1·21 to 1·76), with inadequate PPE associated with a comparable 31% increase (aHR 1·31, 95% CI: 1·10 to 1·56; **Table 3**).

**Table 3.** Risk of reporting a positive test for COVID-19 according to personal protective equipment (PPE) availability and exposure to COVID-19 patients among frontline healthcare workers.

|  | Personal protective equipment |  |  |
| --- | --- | --- | --- |
|  | Adequate | Reused | Inadequate |
| No. with Event/Person-days | 592/332,901 | 146/80,728 | 157/60,916 |
| Unadjusted HR (95% CI) | 1.0 (ref.) | 1.46 (1.21 to 1.76) | 1.32 (1.10 to 1.57) |
| Multivariate-adjusted HR (95% CI) | 1.0 (ref.) | 1.46 (1.21 to 1.76) | 1.31 (1.10 to 1.56) |
| **<u>Exposure to patients with COVID-19</u>** |  |  |  |
| **None** |  |  |  |
| No. with Event/Person-days | 186/227,654 | 19/37,599 | 48/35,159 |
| Unadjusted HR (95% CI) | 1.0 (ref.) | 0.96 (0.60 to 1.55) | 1.53 (1.11 to 2.11) |
| Multivariate-adjusted HR (95% CI) | 1.0 (ref.) | 0.95 (0.59 to 1.54) | 1.52 (1.10 to 2.09) |
| **Suspected COVID-19 patients** |  |  |  |
| No. with Event/Person-days | 126/54,676 | 36/19,378 | 26/14,083 |
| Unadjusted HR (95% CI) | 2.40 (1.91 to 3.02) | 3.23 (2.24 to 4.66) | 1.87 (1.24 to 2.83) |
| Multivariate-adjusted HR (95% CI) | 2.39 (1.90 to 3.00) | 3.20 (2.22 to 4.61) | 1.83 (1.21 to 2.78) |
| **Documented COVID-19 patients** |  |  |  |
| No. with Event/Person-days | 280/50,571 | 91/23,751 | 83/11,675 |
| Unadjusted HR (95% CI) | 4.93 (4.07 to 5.97) | 5.12 (3.94 to 6.64) | 5.95 (4.57 to 7.76) |
| Multivariate-adjusted HR (95% CI) | 4.83 (3.99 to 5.85) | 5.06 (3.90 to 6.57) | 5.91 (4.53 to 7.71) |

Abbreviations: CI (confidence interval), HR (hazard ratio)
All models were stratified by 5-year age group, calendar date at study entry, and country.
Multivariate risk factor models were adjusted for sex, history of diabetes, heart disease, lung disease, kidney disease, and current smoking (each yes/no), and body mass index (17-19.9, 20-24.9, 25-29.9, and $\geq 30$ kg/m<sup>2</sup>).

Frontline HCWs with inadequate PPE in direct contact with a documented COVID-19 positive patient had an aHR of 5·91 (95% CI: 4·53 to 7·71) for a positive COVID-19 test compared to those with adequate PPE who were not in contact with suspected or documented COVID-19 patients. The corresponding aHR was 5·06 (95% CI: 3·90 to 6·57) for those reporting reuse of PPE exposed to patients with documented COVID-19 infection. Notably, even among those reporting adequate PPE, the aHR for a positive COVID-19 test was 2·39 (95% CI: 1·90 to 3·00) for those caring for suspected COVID-19 patients and 4.83 (95% CI: 3·99 to 5·85) for those caring for documented COVID-19 patients compared with HCWs who did not care for either group (**Table 3**).

### Workplace location

We examined whether the elevated risk of a positive COVID-19 test differed according to practice location. Compared to the general community, the aHRs for frontline HCWs were 24·3 (95% CI: 21·8 to 27·1) for those working in inpatient settings; 16·2 (95% CI: 13·4 to 19·7) for nursing homes; 11·2 (95% CI: 8·44 to 14·9) for hospital-based clinics; 7·86 (95% CI: 5·63 to 11·0) for home health sites; 6·94 (95% CI: 5·12 to 9·41) for free-standing ambulatory clinics; and 9·52 (95% CI: 7·49 to 12·1) for all others (Table 4). Notably, HCWs in nursing homes were the most frequent (16.9%) to report inadequate supplies of PPE, while inpatient providers reported reusing PPE 23·7% of the time.

**Table 4.**
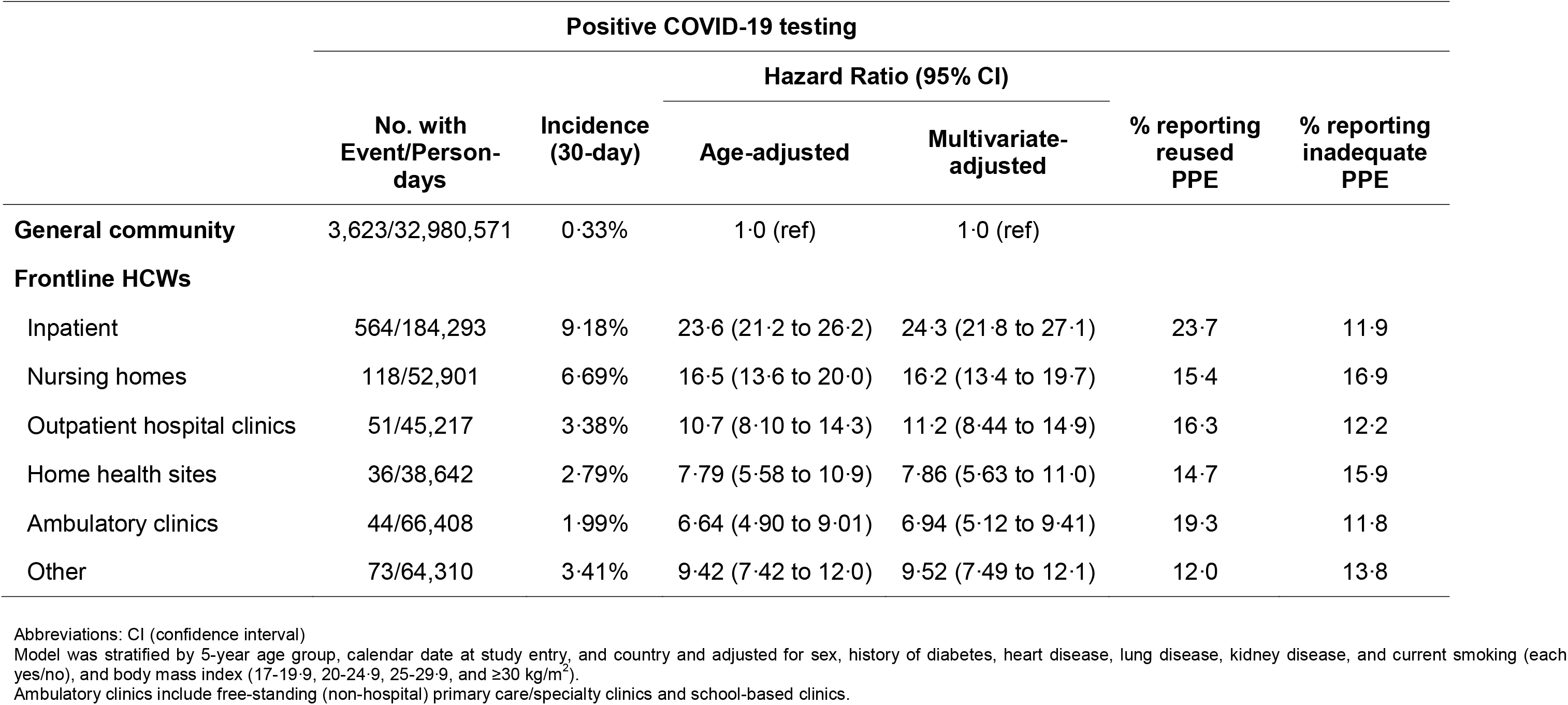
Frontline healthcare workers and risk of testing positive for COVID-19 by site of care delivery.

|  | Positive COVID-19 testing |  |  |  |  |  |
| --- | --- | --- | --- | --- | --- | --- |
|  | No. with Event/Person-days | Incidence (30-day) | Hazard Ratio (95% CI) |  | % reporting reused PPE | % reporting inadequate PPE |
|  |  |  | Age-adjusted | Multivariate-adjusted |  |  |
| **General community** | 3,623/32,980,571 | 0.33% | 1.0 (ref) | 1.0 (ref) |  |  |
| **Frontline HCWs** |  |  |  |  |  |  |
| Inpatient | 564/184,293 | 9.18% | 23.6 (21.2 to 26.2) | 24.3 (21.8 to 27.1) | 23.7 | 11.9 |
| Nursing homes | 118/52,901 | 6.69% | 16.5 (13.6 to 20.0) | 16.2 (13.4 to 19.7) | 15.4 | 16.9 |
| Outpatient hospital clinics | 51/45,217 | 3.38% | 10.7 (8.10 to 14.3) | 11.2 (8.44 to 14.9) | 16.3 | 12.2 |
| Home health sites | 36/38,642 | 2.79% | 7.79 (5.58 to 10.9) | 7.86 (5.63 to 11.0) | 14.7 | 15.9 |
| Ambulatory clinics | 44/66,408 | 1.99% | 6.64 (4.90 to 9.01) | 6.94 (5.12 to 9.41) | 19.3 | 11.8 |
| Other | 73/64,310 | 3.41% | 9.42 (7.42 to 12.0) | 9.52 (7.49 to 12.1) | 12.0 | 13.8 |

Abbreviations: CI (confidence interval)
Model was stratified by 5-year age group, calendar date at study entry, and country and adjusted for sex, history of diabetes, heart disease, lung disease, kidney disease, and current smoking (each yes/no), and body mass index (17-19.9, 20-24.9, 25-29.9, and $\geq 30$ kg/m<sup>2</sup>).
Ambulatory clinics include free-standing (non-hospital) primary care/specialty clinics and school-based clinics.

## DISCUSSION

Among 2,135,190 participants in the U.K. and U.S. assessed between March 24, 2020 and April 23, 2020, we found that frontline HCWs had up to a 12-fold increased risk of reporting a positive COVID-19 test and predicted COVID-19 infection compared to members of the general community, even after accounting for other risk. The risk appeared higher in the U.K. compared with the U.S. Among frontline HCWs, reuse of PPE or inadequate PPE, which could indicate inadequate supply and/or quality, was associated with a subsequent 31-46% increased risk of COVID-19. Although HCWs caring for COVID-19 patients who reported inadequate PPE had the highest risk, an increased susceptibility to infection was evident even among those reporting adequate PPE. Frontline HCWs who worked in inpatient settings (where providers most frequently reported PPE reuse) and nursing homes (where providers most frequently reported inadequate PPE) had the greatest risk.

Our population-scale findings may help provide greater context for regional and national reports from U.S. public health authorities based on a limited cross section of data suggesting 10-20% of documented COVID-19 infections occur among HCWs.^5-7^ Our results offer individual-level data additionally accounting for community or workplace risk factors that complement these limited reports by providing a more precise assessment of the magnitude of increased risk among HCWs during the initial phases of this pandemic in both the U.K. and U.S. Taken in the context of the requirement for testing to establish a COVID-19 diagnosis, our complementary results based both on reporting a positive test for COVID-19 or predicted COVID infection offer a better range of estimates of the true risk of infection experienced by frontline HCWs in patient-facing roles.

We also offer compelling evidence that sufficient availability and/or quality of PPE reduces the risk of COVID-19 infection, but reuse of PPE or inadequate PPE may confer comparably elevated risk, the first study to specifically explore PPE reuse.^17^ The greater risk associated with reuse of PPE could be related to self-contamination during repeated donning and doffing or breakdown of materials due to extended wear. Of note, during the time period of this study, disinfection protocols prior to reuse of PPE were not widely available in the U.S. or U.K.^12-14^ Thus, these results should be not extended to reflect risk of reusing PPE after such disinfection, which has been implemented in a variety of care settings over the last several weeks. A global assessment of the adequacy of the supply chain for PPE should be a part of the deliberate and informed decision making needed prior to lifting stay-at-home orders.

However, even with adequate PPE, HCWs who cared for patients with COVID-19 remained at elevated risk, highlighting the importance of ensuring PPE quality and availability, but also other aspects of appropriate usage, including correct donning and doffing and clinical situation (practice location). In addition, the apparent lack of complete protection against acquiring COVID-19 infection through adequate PPE suggests that additional risk mitigation strategies require further investigation. Moreover, these data underscore the possibility for HCWs to perpetuate infections or contribute to community spread, especially when asymptomatic or mildly symptomatic, and justify calls to increase testing to reduce hospital-based transmission.^4^

Notably, we found a significant difference in the magnitude of aHRs for HCWs in the U.K. compared with the U.S. This may be due to country or region-specific variation in population density, socioeconomic deprivation, overall availability or quality of PPE, and type of healthcare settings that require further investigation. These results may also reflect differences in access to testing among HCWs compared to the general community in the U.K. compared with the U.S. However, in secondary analyses using inverse probability-weighted Cox modeling adjusting for the probability of receiving a test, we also found that HCWs in the U.K. were at higher risk of reporting a positive test. Furthermore, using a symptom-based classification model for predicted COVID-19 positivity, HCWs were at greater risk of developing symptoms predictive of COVID-19, which does not reflect access to testing. Thus, the higher risk observed in the U.K. may reflect a higher infection rate due to differences in the quality and appropriate usage of PPE across practice settings^18^ or country-specific differences in PPE recommendations for HCWs or the general public (e.g. cloth face coverings).^19,20^ Ideally, we would assess COVID-19 risk within a population which has undergone uniform screening. However, the current shortage of PCR-based testing kits does not make such an approach feasible but may justify targeted screening of frontline HCWs.^4,21^ Future studies using serologic testing to ascertain COVID-19 infection will require assessments of test performance and the ability to distinguish recent or active infection from past exposure.

Our results are supported by historical data during similar infectious disease outbreaks. During the Ebola crisis, a disease with a comparable reproduction number (e.g. the R_0_ measure of new cases generated from one individual), HCWs comprised 3·9% of all cases, 21-to-32-times greater than the general public.^22^ During the severe acute respiratory syndrome coronavirus (SARS-CoV or SARS) epidemic, HCWs comprised 20-40% of cases,^23-25^ and inadequate PPE availability was associated with increased risk among HCWs.^25^ The experience with influenza A virus subtype H1N1 reaffirmed the importance of PPE^26^ and showed much higher infection rates among HCWs in dedicated infection containment units.^27^

The strengths of this study include the use of a mobile application to rapidly collect prospective data from a large multinational cohort in real-time, which offers immediately actionable risk estimates to inform the public health response to an ongoing pandemic.^28^ By recruiting participants through existing cohort studies (https://www.monganinstitute.org/cope-consortium),^29^ these results also provide proof-of-concept of the feasibility of leveraging existing infrastructure and engaged participants to address a key knowledge gap. Second, we collected information from participants initially free of a positive COVID-19 test, which offered an opportunity to prospectively assess risk factors for incident infection with minimal recall bias. Third, our study design documented initial onset of symptoms, which minimizes biases related to capturing only more severe cases through hospitalization records or death reports. Finally, we collected information on a wide range of known/suspected risk factors for COVID-19 infection generally not available in existing registries or population-scale surveillance efforts.

We acknowledge several limitations. First, full details of some exposures were limited to ensure our survey was brief. For example, we did not ask about specific occupations, experience level, type of PPE used (e.g. surgical masks, respirators, or powered air purifying respirators), receipt of PPE training (e.g. mask fit-testing or donning and doffing), frequency of exposure to patients with COVID-19 infection or aerosolizing procedures (e.g. endoscopy or intubation). Second, our findings are based on self-report. However, alternative exposure measures, such as PPE supply, or assessment of additional outcomes in such a large cohort would have been difficult to collect in a timely manner within the context of a fast-moving pandemic. In future studies, linkage to other sources (e.g. electronic health records) may be possible. Third, our cohort is not a random sampling of the population. Although this limitation is inherent to any study requiring voluntary provision of health information, we acknowledge that data collection through smartphone adoption has comparatively lower penetrance among certain socioeconomic groups, as well as older adults, despite being used by 81% of the U.S. adult population.^30^ In future studies, we plan more targeted outreach of underrepresented populations, as well as additional collection instruments (web or phone surveys) that may be more accessible. Our primary outcome was based on the report of a positive COVID-19 test. During the study period, this would generally reflect a positive PCR-based swab, which should be moderately specific, as opposed to antibody testing, which was not widely available. However, any misclassification of positive testing should be non-differential according to occupation.

In conclusion, within a large population-based sample of 2,135,190 individuals in the U.S. and U.K., we observed a significantly increased risk of COVID-19 infection among frontline HCWs compared to the general community. This risk is greatest among individuals in direct contact with COVID-19 patients who report inadequate PPE availability or were required to reuse PPE, supporting the importance of providing sufficient high-quality PPE. However, because infection risk remained elevated even with adequate PPE, our results suggest the need to ensure their proper use as well as adherence to other infection control measures. Further studies exploring modifiable and non-modifiable risk factors for HCW-related COVID-19 infection are urgently needed.

## Data Availability

The datasets and code used for the current study are available from the corresponding author on reasonable request. Example code used for the analysis is available on GitHub.

https://github.com/epimath/cm-dag

## Acknowledgements

We would like to thank the more than 2·9 million contributing citizen researchers who have downloaded the COVID Symptom Study, including participants of cohort studies within the Coronavirus Pandemic Epidemiology (COPE) Consortium (**Suppl. Table 1**). We thank the investigators of the cohort studies enrolled in the COPE Consortium for their assistance in disseminating the COVID Symptom Tracker to their study participants, as well as the MGH Clinical and Translational Epidemiology Unit (CTEU) Clinical Research Coordination team. We also thank the staff of Zoe Global Ltd for providing technical support for the app. We also thank Stand Up to Cancer for their assistance in media and social media outreach.

## Author contributions

Study concept and design (LHN, DAD, JW, SO, CJS, TDS, ATC); acquisition of data (LHN, DAD, JW, SO, RD, JCP, CJS, TDS, ATC); data analysis (LHN, DAD, ADJ, CGG, WM, RSM, DRS, CHL, SK, MS, MKG); initial drafting of the manuscript (LHN, DAD, ATC); interpretation of data and critical revision of the manuscript (all authors); study supervision (SO, CJS, TDS, ATC).

## Data availability

Data collected in the app is being shared with other health researchers through the NHS-funded Health Data Research U.K. (HDRUK)/SAIL consortium, housed in the U.K. Secure Research Platform (UKSeRP) in Swansea. Anonymized data is available to be shared with bonafide HDRUK researchers according to their protocols in the public interest (https://healthdatagateway.org/detail/9b604483-9cdc-41b2-b82c-14ee3dd705f6). U.S. investigators are encouraged to coordinate data requests through the COPE Consortium (www.monganinstitute.org/cope-consortium). Data updates can be found on https://covid.joinzoe.com.

## References

1. Anderson RM, Heesterbeek H, Klinkenberg D, Hollingsworth TD. How will country-based mitigation measures influence the course of the COVID-19 epidemic? Lancet (London, England) 2020; 395(10228): 931–4.

2. Johns Hopkins University of Medicine Coronavirus Resource Center. 2020. https://coronavirus.jhu.edu/us-map (accessed 5/6/2020 2020).

3. The L. COVID-19: protecting health-care workers. Lancet (London, England) 2020; 395(10228): 922.

4. Black JRM, Bailey C, Przewrocka J, Dijkstra KK, Swanton C. COVID-19: the case for health-care worker screening to prevent hospital transmission. Lancet (London, England) 2020; 395(10234): 1418–20.

5. California Department of Public Health Latest Covid-19 Facts. 2020. https://www.cdph.ca.gov/Programs/OPA/Pages/NR20-065.aspx (accessed May 1, 2020 2020).

6. Wisconsin Department of Health Services Covid-19: Wisconsin Cases. 2020. https://www.dhs.wisconsin.gov/covid-19/cases.htm (accessed May 2, 2020 2020).

7. Team CC-R. Characteristics of Health Care Personnel with COVID-19 - United States, February 12-April 9, 2020. MMWR Morb Mortal Wkly Rep 2020; 69(15): 477–81.

8. Lazzerini M, Putoto G. COVID-19 in Italy: momentous decisions and many uncertainties. Lancet Glob Health 2020.

9. Verbeek JH, Rajamaki B, Ijaz S, et al. Personal protective equipment for preventing highly infectious diseases due to exposure to contaminated body fluids in healthcare staff. Cochrane Database Syst Rev 2020; 4: CD011621.

10. Center for Disease Control and Prevention: Interim Infection Prevention and Control Recommendations for Patients with Suspected of Confirmed Coronavirus Disease 2019 (COVID-19) in Health Care Settings. https://www.cdc.gov/coronavirus/2019-ncov/hcp/infection-control-recommendations.html (accessed April 13, 2020.

11. England PH. Guidance: COVID-19 personal protective equipment (PPE). https://www.gov.uk/government/publications/wuhan-novel-coronavirus-infection-prevention-and-control/covid-19-personal-protective-equipment-ppe#summary-of-ppe-recommendations-for-health-and-social-care-workers (accessed 5/20/2020 2020).

12. Fischer R, Morris DH, van Doremalen N, et al. Assessment of N95 respirator decontamination and re-use for SARS-CoV-2. *medRxiv* 2020: 2020.04.11.20062018.

13. Schwartz A, Stiegel M, Greeson N, et al. Decontamination and Reuse of N95 Respirators with Hydrogen Peroxide Vapor to Address Worldwide Personal Protective Equipment Shortages During the SARS-CoV-2 (COVID-19) Pandemic. Applied Biosafety 2020: 1535676020919932.

14. Livingston E, Desai A, Berkwits M. Sourcing Personal Protective Equipment During the COVID-19 Pandemic. Jama 2020.

15. Drew DA, Nguyen LH, Steves CJ, et al. Rapid implementation of mobile technology for real-time epidemiology of COVID-19. Science 2020: eabc0473.

16. Menni C, Valdes A, Freidin M, et al. Loss of smell and taste in combination with other symptoms is a potential predictor of COVID-19 infection (in press) Nature Medicine 2020.

17. Chou R, Dana T, Buckley DI, Selph S, Fu R, Totten AM. Epidemiology of and Risk Factors for Coronavirus Infection in Health Care Workers. Ann Intern Med 2020.

18. Fifth of frontline doctors complain of unusable coronavirus PPE. The Times. 2020.

19. Thomas JP, Srinivasan A, Wickramarachchi CS, Dhesi PK, Hung YM, Kamath AV. Evaluating the national PPE guidance for NHS healthcare workers during the COVID-19 pandemic. Clin Med (Lond) 2020.

20. Cheng KK, Lam TH, Leung CC. Wearing face masks in the community during the COVID-19 pandemic: altruism and solidarity. Lancet (London, England) 2020.

21. Treibel TA, Manisty C, Burton M, et al. COVID-19: PCR screening of asymptomatic health-care workers at London hospital. Lancet (London, England) 2020.

22. Houlihan CF, McGowan CR, Dicks S, et al. Ebola exposure, illness experience, and Ebola antibody prevalence in international responders to the West African Ebola epidemic 2014–2016: A cross-sectional study. PLoS Med 2017; 14(5): e1002300.

23. Koh D, Lim MK, Chia SE. SARS: health care work can be hazardous to health. Occup Med (Lond) 2003; 53(4): 241–3.

24. Seto WH, Tsang D, Yung RW, et al. Effectiveness of precautions against droplets and contact in prevention of nosocomial transmission of severe acute respiratory syndrome (SARS). Lancet (London, England) 2003; 361(9368): 1519–20.

25. Lau JT, Fung KS, Wong TW, et al. SARS transmission among hospital workers in Hong Kong. Emerg Infect Dis 2004; 10(2): 280–6.

26. Marshall C, Kelso A, McBryde E, et al. Pandemic (H1N1) 2009 risk for frontline health care workers. Emerg Infect Dis 2011; 17(6): 1000–6.

27. Chen MI, Lee VJ, Barr I, et al. Risk factors for pandemic (H1N1) 2009 virus seroconversion among hospital staff, Singapore. Emerg Infect Dis 2010; 16(10): 1554–61.

28. Brownstein JS, Freifeld CC, Madoff LC. Digital disease detection--harnessing the Web for public health surveillance. N Engl J Med 2009; 360(21): 2153–5, 7.

29. Chan AT, Drew DA, Nguyen LH, et al. The COronavirus Pandemic Epidemiology (COPE) Consortium: A Call to Action. Cancer Epidemiology, Biomarkers & Prevention 2020: cebp.0606.2020.

30. Pew Research Center for Internet & Technology: Mobile Fact Sheet. 2020. https://www.pewresearch.org/internet/fact-sheet/mobile/ (accessed April 27, 2020 2020).

